# A neutrophil activation signature predicts critical illness and mortality in COVID-19

**DOI:** 10.1101/2020.09.01.20183897

**Authors:** Matthew L. Meizlish, Alexander B. Pine, Jason D. Bishai, George Goshua, Emily R. Nadelmann, Michael Simonov, C-Hong Chang, Hanming Zhang, Marcus Shallow, Parveen Bahel, Kent Owusu, Yu Yamamoto, Tanima Arora, Deepak S. Atri, Amisha Patel, Rana Gbyli, Jennifer Kwan, Christine H. Won, Charles Dela Cruz, Christina Price, Jonathan Koff, Brett A. King, Henry M. Rinder, F. Perry Wilson, John Hwa, Stephanie Halene, William Damsky, David van Dijk, Alfred I. Lee, Hyung J. Chun

## Abstract

Pathologic immune hyperactivation is emerging as a key feature of critical illness in COVID-19, but the mechanisms involved remain poorly understood. We carried out proteomic profiling of plasma from cross-sectional and longitudinal cohorts of hospitalized patients with COVID-19 and analyzed clinical data from our health system database of over 3,300 patients. Using a machine learning algorithm, we identified a prominent signature of neutrophil activation, including resistin, lipocalin-2, HGF, IL-8, and G-CSF, as the strongest predictors of critical illness. Neutrophil activation was present on the first day of hospitalization in patients who would only later require transfer to the intensive care unit, thus preceding the onset of critical illness and predicting increased mortality. In the health system database, early elevations in developing and mature neutrophil counts also predicted higher mortality rates. Altogether, we define an essential role for neutrophil activation in the pathogenesis of severe COVID-19 and identify molecular neutrophil markers that distinguish patients at risk of future clinical decompensation.

## Main

As the death toll from COVID-19 reaches over 830,000 worldwide, it remains a pressing concern to understand how the disease causes such a wide spectrum of clinical outcomes. For the majority of patients, COVID-19 manifests as an upper respiratory tract infection that is self-limited. However, the progression of COVID-19 in a large subset of patients to respiratory distress, multi-organ failure, and death has resulted in an enormous global impact. A number of studies to date have highlighted an important role for monocytes and macrophages in severe COVID-19,^1,2^ but our knowledge of the immunological drivers of critical illness are otherwise limited.

To achieve a deeper understanding of the immunological phenotypes of COVID-19 across the spectrum of disease severity, we performed proteomic profiling of blood obtained from hospitalized patients with COVID-19 and used a machine learning algorithm to define the biomarkers that best discriminate between critically ill patients and those with more mild disease. We discovered a unique neutrophil activation signature composed of neutrophil activators (G-CSF, IL-8) and effectors (resistin (RETN), lipocalin-2 (LCN2), and hepatocyte growth factor (HGF)), which had the greatest power of all measured biomarkers to identify critically ill patients. The effector proteins strongly correlated with absolute neutrophil count and were highly transcriptionally enriched in a developing neutrophil population that was recently identified specifically in critically ill COVID-19 patients.^3,4^ To determine whether this neutrophil activation signature precedes the onset of critical illness, we performed longitudinal plasma analyses beginning on day 1 of patients’ hospitalization. We discovered that elevated neutrophil biomarkers at the time of hospital admission identified those patients who would later progress to critical illness and predicted increased in-hospital mortality. Finally, to determine whether the mechanistic implications of this study are generalizable, we analyzed clinical data from a cohort of over 3,300 patients and found that, at the earliest blood draw during hospitalization, high numbers of developing and mature neutrophils detected in patients’ blood also predicted increased mortality. This study identifies neutrophil activation as a defining feature of severe COVID-19 that occurs prior to the onset of critical illness, implicating neutrophils as a central player in the pathogenesis of severe COVID-19 and highlighting opportunities for clinical prediction and therapeutic intervention.

## Results

Forty-nine adult patients (40 in the medical intensive care unit (ICU) and 9 in non-ICU units), as well as 13 non-COVID-19, non-hospitalized controls, were included in our initial ‘cross-sectional’ cohort (**Table 1a**). We performed multiplexed biomarker profiling on plasma obtained during the course of patients’ hospitalization, measuring concentrations of 78 circulating proteins with immunologic functions (**Figure 1a**, **Supp. Fig. 1**). Principal component analysis (PCA) of this dataset showed separation of the control, non-ICU COVID-19, and ICU COVID-19 samples, indicating that these circulating markers capture a spectrum of illness severity (**Figure 1b**).

**Table 1.**
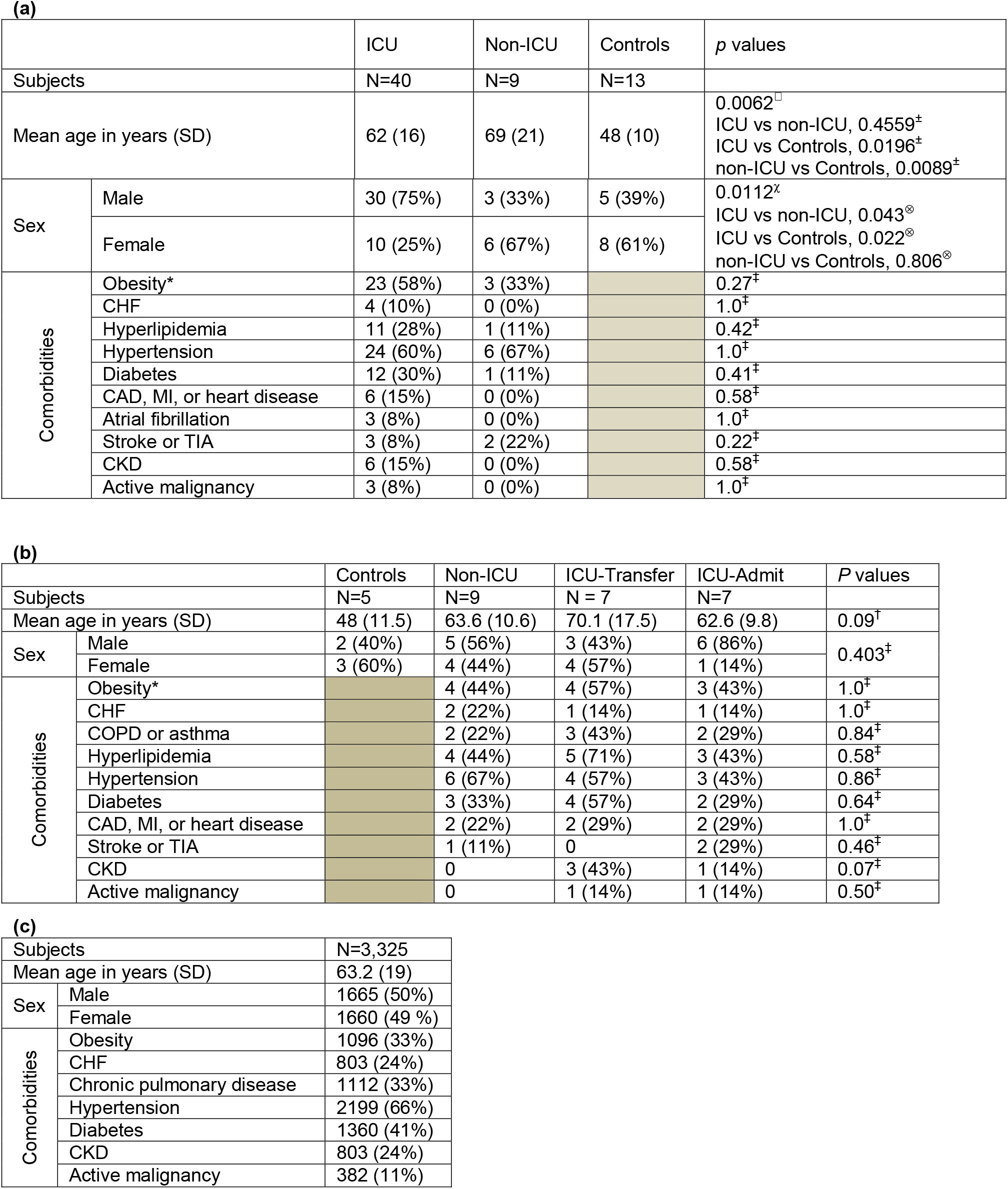
Demographics of patients in the cross-sectional (a), longitudinal (b), and DOM-CovX (c) cohorts. *Obesity is defined as BMI>30. One-way ANOVA with ^±^post-hoc Tukey’s multiple comparisons tests. ^χ^Group-wise Chi^2^ test; ^⊗^individual Chi^2^ tests. ^†^Kruskal-Wallis test. ^‡^Fisher’s exact test. Abbreviations: SD, standard deviation; CHF, congestive heart failure; CAD, coronary artery disease; MI, myocardial infarction; TIA, transient ischemic attack, CKD, chronic kidney disease

|  |  | ICU | Non-ICU | Controls | p values |
| --- | --- | --- | --- | --- | --- |
| Subjects |  | N=40 | N=9 | N=13 |  |
| Mean age in years (SD) |  | 62 (16) | 69 (21) | 48 (10) | 0.0062<sup>□</sup><br>ICU vs non-ICU, 0.4559<sup>+</sup><br>ICU vs Controls, 0.0196<sup>+</sup><br>non-ICU vs Controls, 0.0089<sup>+</sup> |
| Sex | Male | 30 (75%) | 3 (33%) | 5 (39%) | 0.0112<sup>x</sup><br>ICU vs non-ICU, 0.043<sup>®</sup><br>ICU vs Controls, 0.022<sup>®</sup><br>non-ICU vs Controls, 0.806<sup>®</sup> |
|  | Female | 10 (25%) | 6 (67%) | 8 (61%) |  |
| Comorbidities | Obesity* | 23 (58%) | 3 (33%) |  | 0.27<sup>†</sup> |
|  | CHF | 4 (10%) | 0 (0%) |  | 1.0<sup>‡</sup> |
|  | Hyperlipidemia | 11 (28%) | 1 (11%) |  | 0.42<sup>‡</sup> |
|  | Hypertension | 24 (60%) | 6 (67%) |  | 1.0<sup>‡</sup> |
|  | Diabetes | 12 (30%) | 1 (11%) |  | 0.41<sup>‡</sup> |
|  | CAD, MI, or heart disease | 6 (15%) | 0 (0%) |  | 0.58<sup>‡</sup> |
|  | Atrial fibrillation | 3 (8%) | 0 (0%) |  | 1.0<sup>‡</sup> |
|  | Stroke or TIA | 3 (8%) | 2 (22%) |  | 0.22<sup>‡</sup> |
|  | CKD | 6 (15%) | 0 (0%) |  | 0.58<sup>‡</sup> |
|  | Active malignancy | 3 (8%) | 0 (0%) |  | 1.0<sup>‡</sup> |

(b)
|  |  | Controls | Non-ICU | ICU-Transfer | ICU-Admit | P values |
| --- | --- | --- | --- | --- | --- | --- |
| Subjects |  | N=5 | N=9 | N = 7 | N=7 |  |
| Mean age in years (SD) |  | 48 (11.5) | 63.6 (10.6) | 70.1 (17.5) | 62.6 (9.8) | 0.09<sup>†</sup> |
| Sex | Male | 2 (40%) | 5 (56%) | 3 (43%) | 6 (86%) | 0.403<sup>‡</sup> |
|  | Female | 3 (60%) | 4 (44%) | 4 (57%) | 1 (14%) |  |
| Comorbidities | Obesity* |  | 4 (44%) | 4 (57%) | 3 (43%) | 1.0<sup>‡</sup> |
|  | CHF |  | 2 (22%) | 1 (14%) | 1 (14%) | 1.0<sup>‡</sup> |
|  | COPD or asthma |  | 2 (22%) | 3 (43%) | 2 (29%) | 0.84<sup>‡</sup> |
|  | Hyperlipidemia |  | 4 (44%) | 5 (71%) | 3 (43%) | 0.58<sup>‡</sup> |
|  | Hypertension |  | 6 (67%) | 4 (57%) | 3 (43%) | 0.86<sup>‡</sup> |
|  | Diabetes |  | 3 (33%) | 4 (57%) | 2 (29%) | 0.64<sup>‡</sup> |
|  | CAD, MI, or heart disease |  | 2 (22%) | 2 (29%) | 2 (29%) | 1.0<sup>‡</sup> |
|  | Stroke or TIA |  | 1 (11%) | 0 | 2 (29%) | 0.46<sup>‡</sup> |
|  | CKD |  | 0 | 3 (43%) | 1 (14%) | 0.07<sup>‡</sup> |
|  | Active malignancy |  | 0 | 1 (14%) | 1 (14%) | 0.50<sup>‡</sup> |

(c)
|  |  |  |
| --- | --- | --- |
| Subjects |  | N=3,325 |
| Mean age in years (SD) |  | 63.2 (19) |
| Sex | Male | 1665 (50%) |
|  | Female | 1660 (49 %) |
| Comorbidities | Obesity | 1096 (33%) |
|  | CHF | 803 (24%) |
|  | Chronic pulmonary disease | 1112 (33%) |
|  | Hypertension | 2199 (66%) |
|  | Diabetes | 1360 (41%) |
|  | CKD | 803 (24%) |
|  | Active malignancy | 382 (11%) |

**Figure 1.**
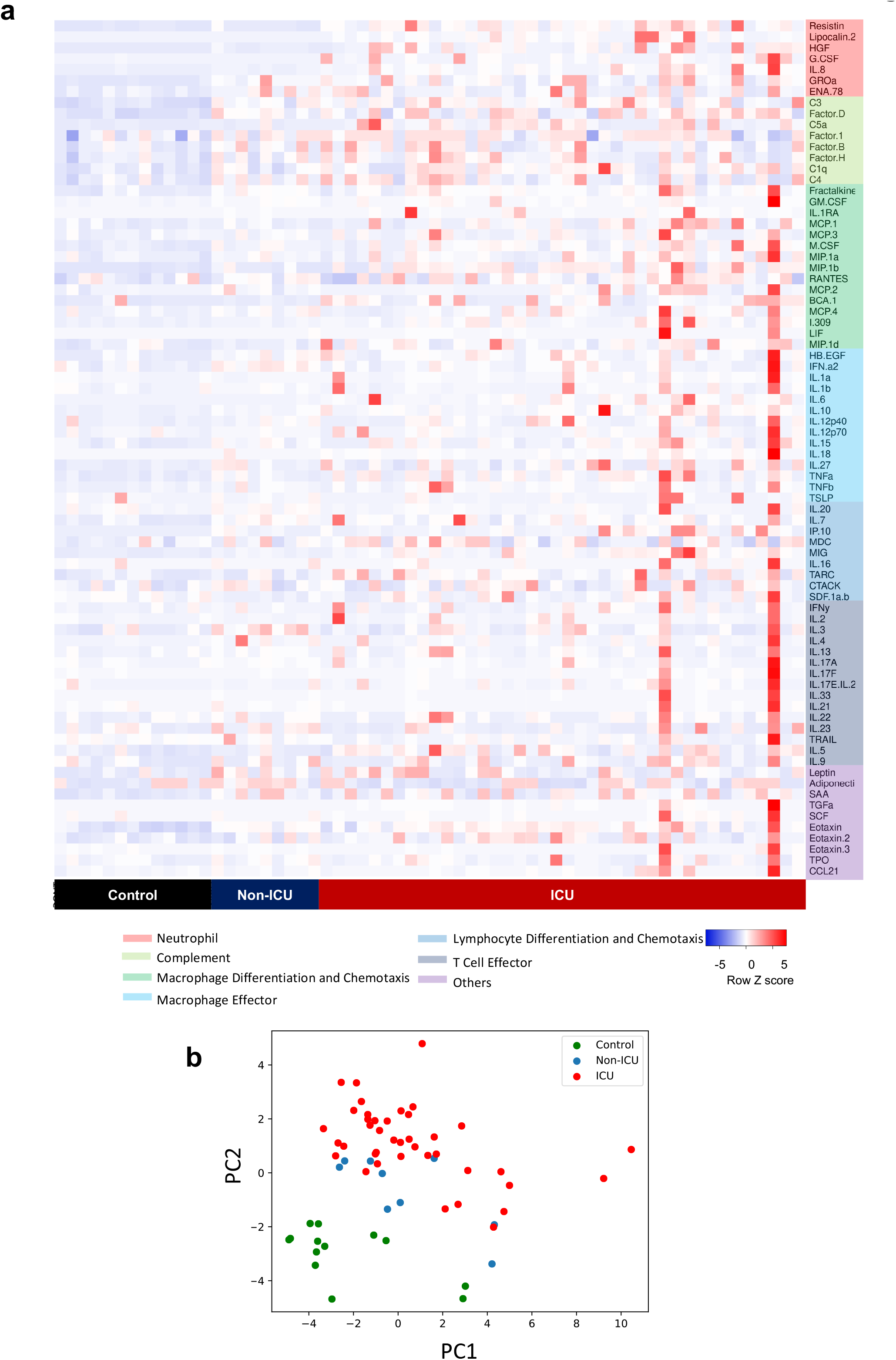
Circulating biomarkers separate COVID-19 patients according to disease severity. (a) Heatmap of proteomic data from the cross-sectional cohort, indicating relative protein levels detected in each subject (columns) for all biomarkers tested (rows). Proteins are categorized by biological function. (b) Visualization of the first two principal components (PC) of a principal component analysis of all biomarker data for each subject.

To assess if this profile of plasma biomarkers could distinguish between critically ill and non-critically ill patients, we applied a random forest machine learning prediction model. The model was trained on data from 2/3 of the subjects, and its ability to predict ICU status was tested on the remaining 1/3 of the subjects. The model accurately identified 14 out of 14 patients in the ICU and 7 out of 7 subjects not in the ICU (**Figure 2a**). To avoid introducing bias based on treatment, the random forest model and PCA analyses excluded IL-6 because most (38/40) ICU patients received tocilizumab, which is known to increase IL-6 levels.^5^ These results demonstrate that our proteomic profile provides a highly reliable circulating signature of critical illness in COVID-19.

**Figure 2.**
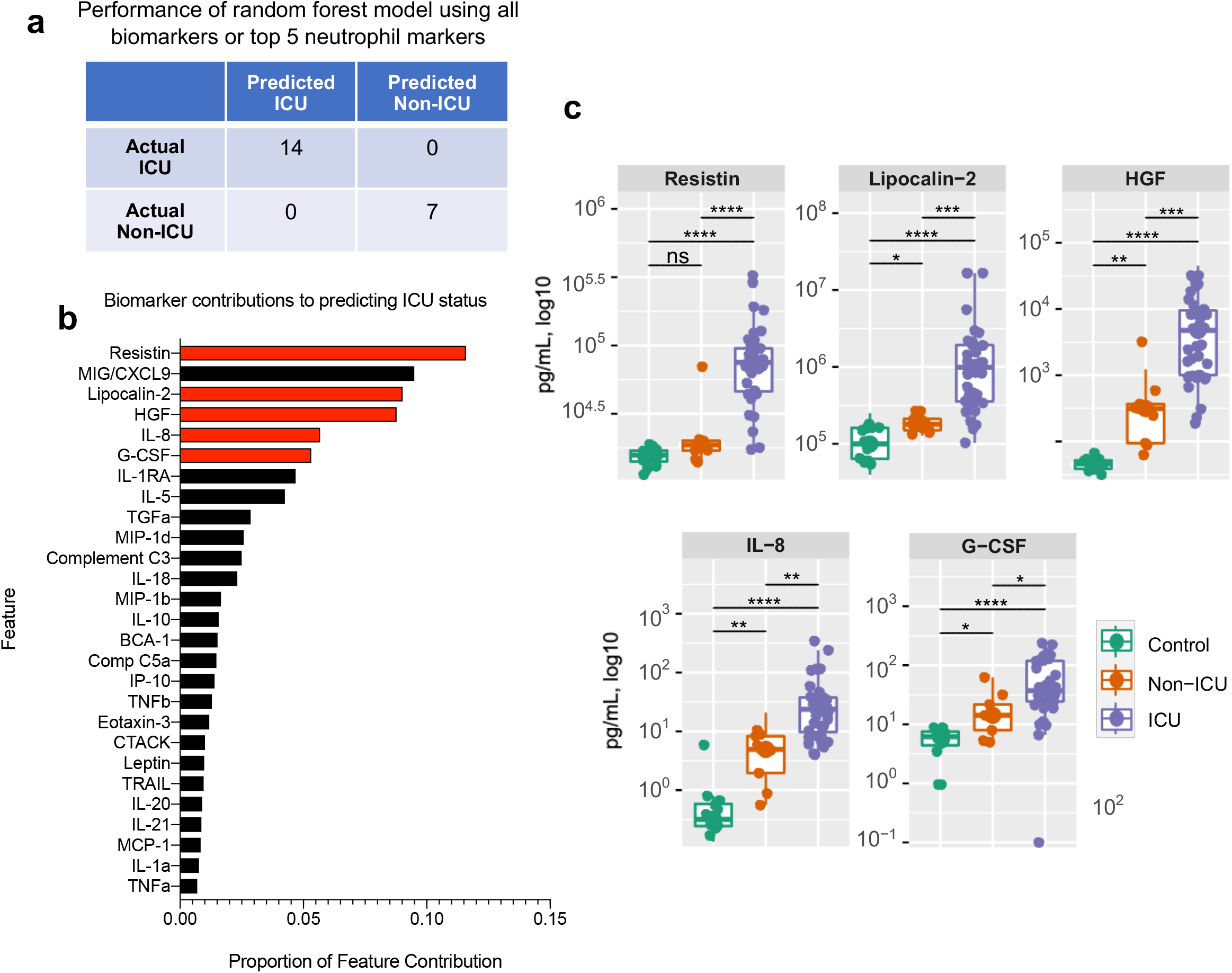
Markers of neutrophil activation accurately identify patients with critical illness. (a) Performance of a random forest (RF) model trained on data from two thirds of the study subjects in the cross-sectional cohort, predicting ICU status for the remaining one third of subjects not included in the training set. Perfect classification as depicted is achieved when using data from all biomarkers or from only the top 5 neutrophil markers (highlighted in red in 2b). (b) Feature importance ranked by proportion of feature contribution to the RF model. Members of the neutrophil activation signature are highlighted in red. (c) Comparisons of circulating levels of neutrophil markers in: 1) controls, 2) non-ICU COVID-19, and 3) ICU COVID-19 patients in the cross-sectional cohort. Asterisks denote statistically significant differences between groups (\**p* < 0.05, \*\**p* < 0.01, \*\*\**p* < 0.001, \*\*\*\**p* < 0.0001). ICU: intensive care unit.

To gain insight into the specific plasma proteins that may affect disease severity, we next examined the importance of each feature in the random forest prediction model. Five of the top six features contributing to the model were proteins related to neutrophil activation (**Figure 2b, 2c**). Three of the top four features were RETN, LCN2, and HGF, each of which is produced by neutrophils, stored in neutrophil ‘specific’ or ‘secondary’ granules, and released upon neutrophil activation.^6-11^ The next two highest ranking features were IL-8 and G-CSF, which stimulate neutrophil chemotaxis and development, respectively.^12,13^

Next, we assessed whether this neutrophil activation signature alone could discriminate between critically ill (ICU) and non-critically ill (non-ICU) patients. We repeated random forest modeling using only the markers of neutrophil activation (RETN, LCN2, HGF, IL-8, and G-CSF) and found that circulating levels of this selective panel also accurately classified all test subjects into ICU and non-ICU categories (**Figure 2a**). Interestingly, each of the neutrophil activation markers showed greater discriminatory power for critical illness than previously described monocyte/macrophage markers (with the exception of MIG/CXCL9) that were also included in our proteomic panel (**Figure 1a, Figure 2b, Supp. Fig. 1**),^14,15^ which suggests an essential role for neutrophils in the development of critical illness associated with COVID-19.

To interrogate the source of neutrophil granule proteins RETN, LCN2, and HGF within our patient cohort, we analyzed the correlation of the absolute neutrophil count (ANC) at the time of blood draw with all of the circulating markers that we had measured. We found that RETN, HGF, and LCN2 were the three proteins that most strongly correlated with ANC (Spearman’s r = 0.633, p < 0.001; 0.555, *p* < 0.001; and 0.492, *p* < 0.0001, respectively) (**Figure 3a**). To gain further insight into the cellular source of these proteins in COVID-19, we analyzed data from a single cell RNA sequencing (scRNAseq) study of peripheral blood mononuclear cells (PBMCs) from patients with COVID-19.^3^ We found that *RETN* and *LCN2* transcripts were highly enriched in a circulating cell population designated as “developing neutrophils” (*p* < 10e^-300^ for each), which was detected almost exclusively in severely ill patients with acute respiratory distress syndrome (ARDS) (**Figure 3b, Supp. Fig. 2**).^3^ Another highly enriched marker (*p* < 10e^-300^) for this population was matrix metallopeptidase 8 *(MMP8)*, or neutrophil collagenase (**Figure 3b**). Notably, constituents of neutrophil granules, like *RETN, LCN2*, and *MMP8*, are transcribed at earlier stages of neutrophil development, packaged into granules, and later released from mature neutrophils upon degranulation.^16^ These observations provide additional support for the conclusion that neutrophils are the primary source of the circulating markers of critical illness in COVID-19 that we identified.

**Figure 3.**
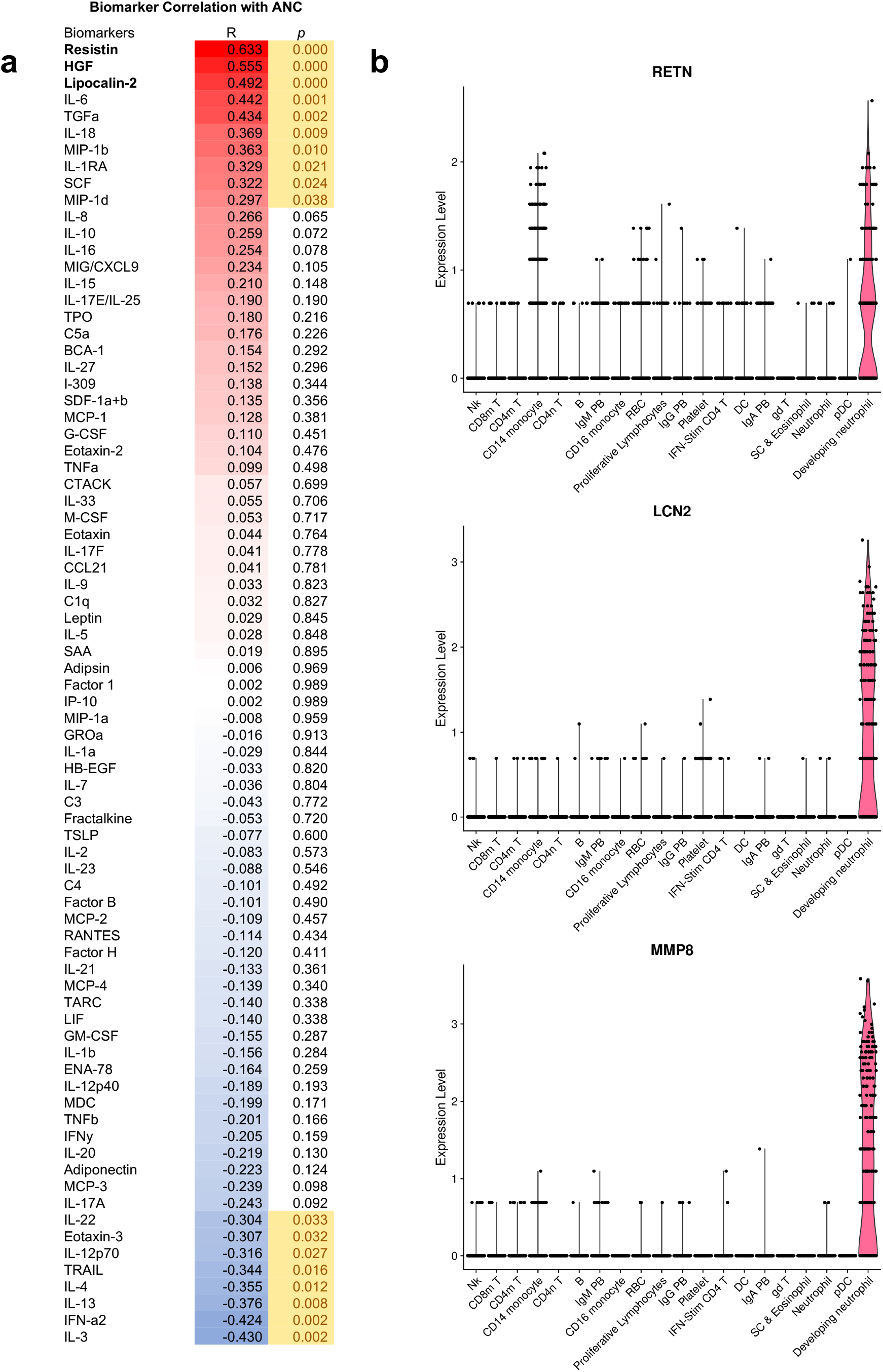
Circulating neutrophil granule proteins are likely derived from neutrophilic source in COVID-19 patients. (a) Correlations of circulating biomarkers with absolute neutrophil count (ANC) in the cross-sectional cohort. Neutrophil granule proteins, which show the highest correlation with ANC, are highlighted. R, Spearman’s rank correlation coefficient; *p*, p-value. (b) Violin plots of resistin (RETN), lipocalin-2 (LCN2), and matrix metallopeptidase 8 (MMP8) mRNA, showing enrichment in a ‘developing neutrophil’ population, based on reanalysis of single-cell RNAseq data published by Wilk et al of peripheral blood mononuclear cells from patients with COVID-19.^3^

In our initial cross-sectional cohort, blood was collected after patients had been hospitalized for a variable length of time (between 2 and 45 days after admission), limiting our capacity to determine whether the observed neutrophil signature was a consequence of critical illness or whether it preceded its onset. To address this, we established a second, longitudinal cohort of patients. Proteomic plasma profiling was conducted on blood samples collected serially starting on day 1 (within 24 hours of hospital admission) from 23 consecutive patients admitted for treatment of confirmed COVID-19 who remained hospitalized for at least 4 days (**Figure 4a, Supp. Fig. 3, 4**). To broaden our assessment of neutrophil activation, we included a panel of matrix metalloproteinases (MMPs) and tissue inhibitor of metalloproteinases (TIMPs), including MMP8, a marker of the ‘developing neutrophil’ population (**Figure 3b**).^17^ Consistent with our findings in the cross-sectional cohort, RETN, HGF, and LCN2 in the day 1 samples of the longitudinal cohort showed strong correlations with ANC (Spearman’s r = 0.76, p < 0.0001; 0.60, p =0.0031; and 0.73, p = 0.0001, respectively), and MMP8 was the protein most highly correlated with ANC (r = 0.86, p < 0.0001), further supporting a neutrophilic source of these proteins in COVID-19 (**Supp. Fig. 5**).

**Figure 4.**
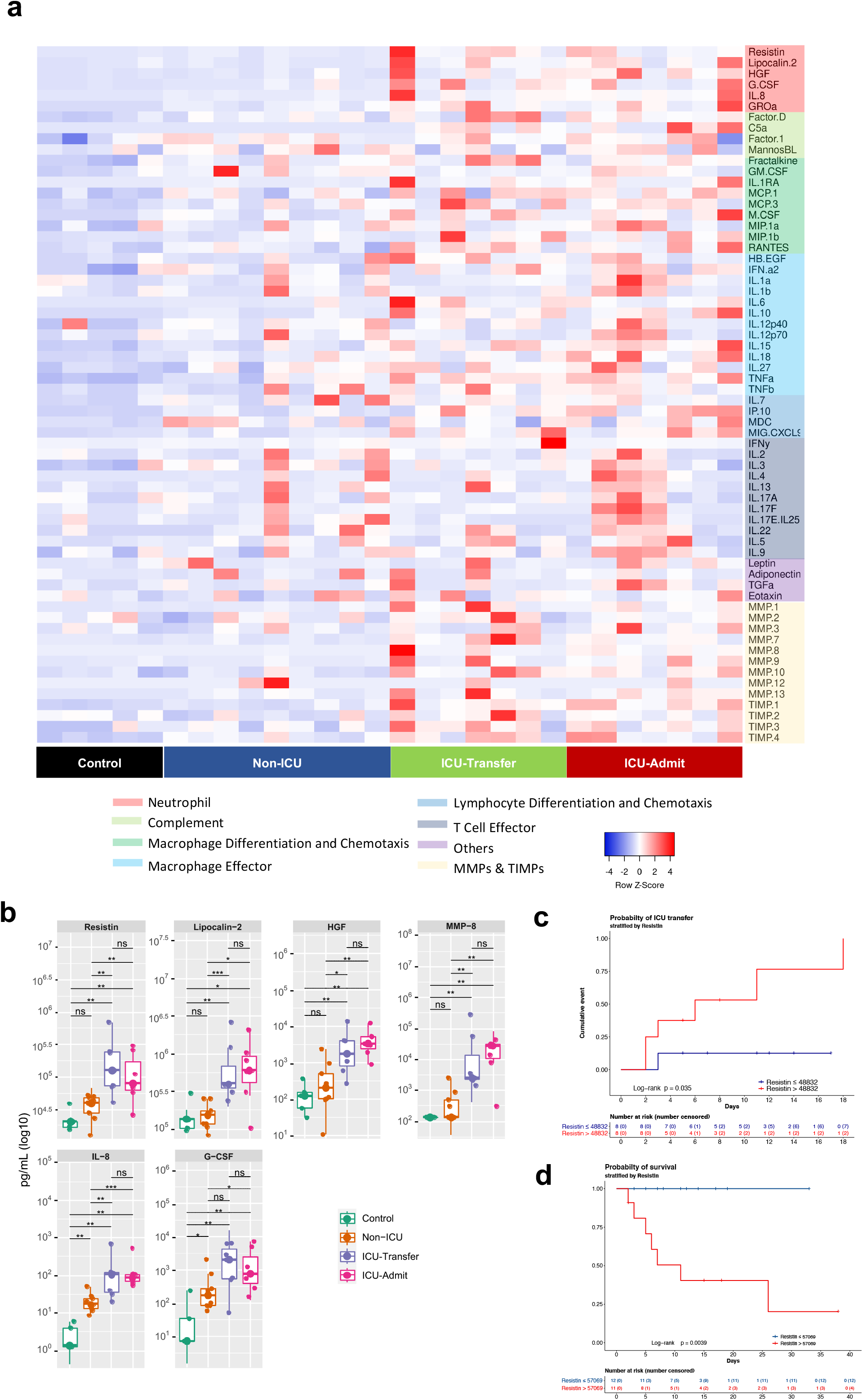
Elevation of the neutrophil activation signature precedes the onset of critical illness. (a) Heatmap indicating relative protein levels detected on day 1 in each subject in the longitudinal cohort (columns) for all biomarkers tested (rows). Proteins are categorized by biological function. (b) Comparisons of circulating levels of neutrophil markers in subjects categorized as: 1) controls, 2) non-ICU, 3) ICU-Transfer, and 4) ICU-Admit. ‘Non-ICU’ indicates patients who remained in a non-ICU unit until discharge; ‘ICU-Transfer’ indicates patients who were admitted to a non-ICU unit and were transferred to an ICU unit during hospitalization; ‘ICU-Admit’ indicates patients who were admitted directly to an ICU unit. Asterisks denote statistically significant differences between groups (\**p* < 0.05, \*\**p* < 0.01, \*\*\**p* < 0.001, *\*\*\*\*p <* 0.0001). (c) Kaplan-Meier curve depicting the likelihood of ICU admission depending on resistin (RETN) levels on day 1, using the median of the group (non-ICU and ICU-Transfer) as the cut-off value. (d) Kaplan-Meier curve depicting the likelihood of survival depending on resistin levels on day 1, using the median of the group (entire longitudinal cohort) as the cut-off value.

Sixteen patients in the longitudinal cohort were initially admitted to lower-acuity non-ICU units, and seven patients were admitted directly to the ICU (‘ICU-Admit’) (**Table 1b**). Of the patients admitted to non-ICU units, nine remained in those units until discharge (‘non-ICU’), while seven required transfer to the ICU during their hospital stay (‘ICU-Transfer’) (six patients were transferred due to worsening respiratory failure (**Supp. Table 1**), and one patient was transferred due to hypotension in the setting of gastrointestinal bleeding). Consistent with our observations in the cross-sectional cohort, ICU-Admit patients had significantly higher levels of all neutrophil activation markers than non-ICU patients on day 1 of hospitalization (**Figure 4b, Supp. Fig. 3b**). As predicted, the same pattern was observed for MMP8 which, like RETN, HGF, and LCN2, is stored and released from secondary granules of neutrophils.^18,19^

Remarkably, the levels of the neutrophil activation markers on day 1 of hospitalization were also significantly elevated in the ICU-Transfer patients, at levels comparable to ICU-Admit patients and well above the levels in non-ICU patients (**Figure 4b**). In both ICU-Transfer and ICU-Admit patients, the neutrophil activation markers remained elevated from the day of admission to day 7 of hospitalization and did not change appreciably over time, while these levels in non-ICU patients remained stably low (**Supp. Fig. 3b**). Thus, despite the fact that patients in the ICU-Transfer group were not critically ill at the time of the day 1 blood draw, evidence of neutrophil activation was already present in these patients. The neutrophil activation signature identified patients who were primed for eventual transfer to the ICU, prior to the onset of critical illness.

Previously reported markers of macrophage activation, including IL-6, IL-10, TNF-α, and MIG/CXCL9 were also elevated in ICU-Admit and ICU-Transfer patients compared to non-ICU patients on day 1 of hospitalization (**Supp. Fig. 4**). Notably, most of these markers (as well as IL-8 and G-CSF) were also significantly elevated in non-ICU patients compared to controls, whereas the neutrophil granule proteins (RETN, LCN2, HGF, MMP8) that directly reflect neutrophil activation were not elevated in non-ICU patients compared to controls and were only significantly different in patients who would go on to develop critical illness (**Figure 4b, Supp. Fig. 4**). Altogether, these data indicate that both neutrophil activation and monocyte/macrophage activation precede the onset of critical illness. They also suggest that neutrophil degranulation may be a more specific feature of severe COVID-19.

To further validate our findings, we conducted additional analyses using RETN, the factor that was most important in distinguishing critical illness in our random forest prediction model. Among patients who did not require direct ICU admission (non-ICU, ICU-Transfer), those with day 1 RETN levels above the median value were much more likely to later require ICU transfer (**Figure 4c**). Moreover, among all patients in this cohort, those with day 1 RETN levels above the median were significantly less likely to survive (**Figure 4d**). In addition to RETN, we found that higher levels on hospital day 1 of LCN2, G-CSF, IL-8, and MMP-8, as well as IL-6, IL-10, TNF-α, IL-1RA, and M-CSF, were significantly associated with mortality (**Supp. Fig. 6**). Meanwhile, we found that the neutrophil to lymphocyte ratio (NLR), which has been reported as a prognostic indicator in COVID-19,^20^ was not significantly different between non-ICU and ICU-Transfer patients on the first day of hospitalization (**Supp. Fig. 7**), suggesting that this measure of cell count ratio may not distinguish critical illness as clearly as the molecular markers of neutrophil activation. Interestingly, neither NLR nor D-dimer, another well-established marker of disease severity in COVID-19, were significantly different between patients who did and did not survive their hospitalization, unlike the neutrophil and macrophage activation markers (**Supp. Fig. 6**).

Lastly, we explored whether these mechanistic insights regarding the role of neutrophils in severe COVID-19 could be validated in a large patient population hospitalized with COVID-19. Using an extensive database of laboratory and clinical data from 3,325 patients admitted to Yale-New Haven Health System who tested positive for SARS-CoV-2 (DOM-CovX cohort; **Table 1c**), we examined the first recorded values of immature granulocyte (IG) and neutrophil counts during the course of their hospitalization. We used these early values in order to avoid potential confounding effects of hospitalization, such as therapeutic interventions and secondary infections. We found that in-hospital mortality was significantly higher among patients with elevated initial IG absolute count (**Figure 5a**), IG percent **(Figure 5b)**, and ANC **(Figure 5c)**. Interestingly, we did not find that initial absolute monocyte count was associated with increased mortality (**Figure 5d**). While our molecular markers of neutrophil activation are not available in this large cohort, these findings confirm that early signs of neutrophil development are associated with future mortality in one of the largest COVID-19 patient cohorts assessed to date.

**Figure 5.**
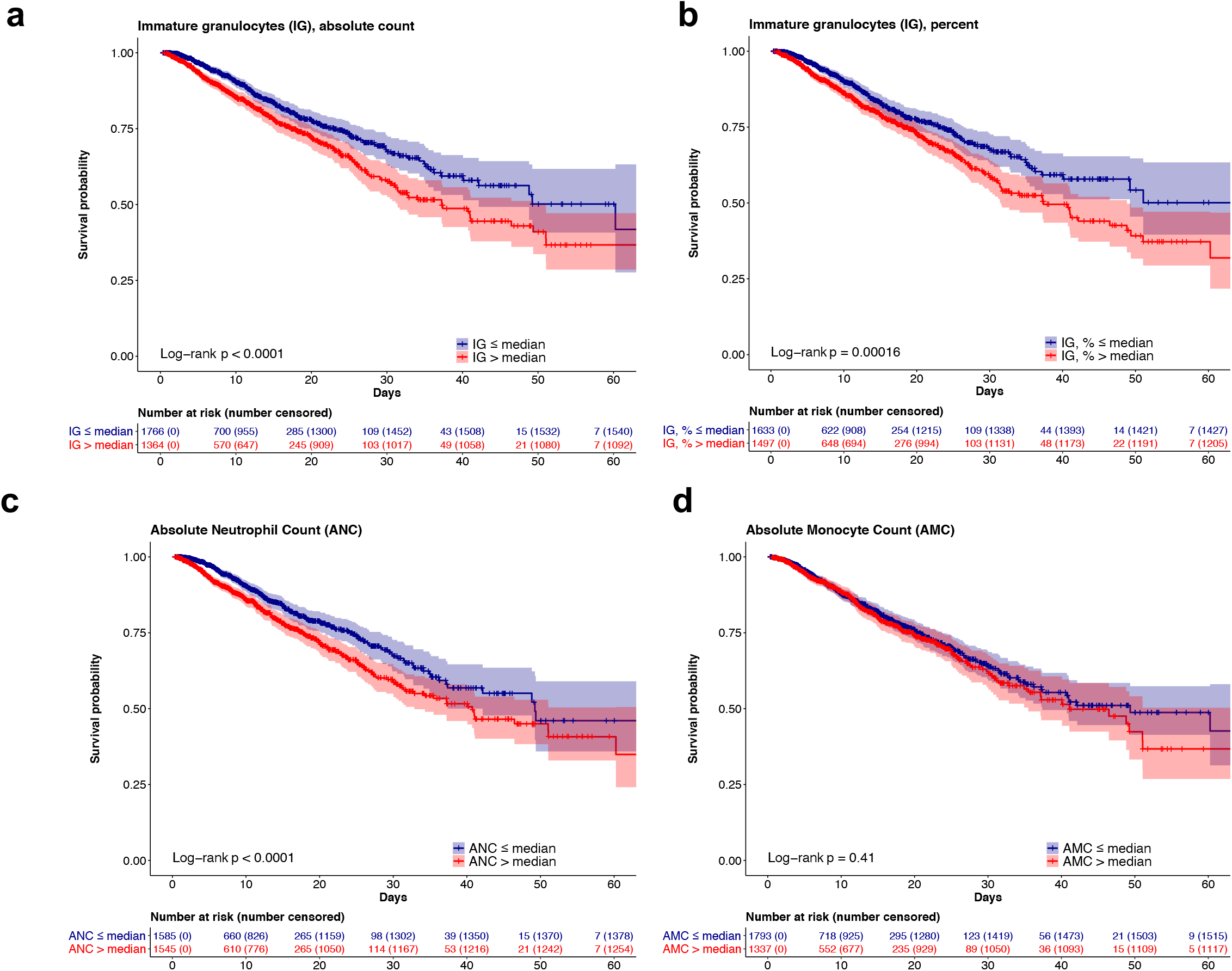
Early elevations in developing and mature neutrophil counts predict increased mortality. Kaplan-Meier curves depicting likelihood of survival based on patients’ first recorded values in the DOM-CovX cohort of (a) absolute immature granulocyte count, (b) immature granulocyte percent, (c) absolute neutrophil count, and (d) absolute monocyte count, using the median value as the cut-off.

## Discussion

Our analyses of multiple hospitalized patient cohorts with COVID-19 reveal a consistent protein-level signature of neutrophil development and activation in critically ill patients with COVID-19. Moreover, we demonstrate for the first time that elevations in circulating markers of neutrophil and macrophage activation precede the onset of critical illness, identifying non-critically ill patients who are at risk of becoming critically ill. Based on our machine learning prediction algorithm, markers of neutrophil activation had greater discriminatory power for detecting critical illness than other cytokines previously shown to be associated with severe COVID-19,^14,15^ most of which were also included in our proteomic profile. Furthermore, in one of the largest cohorts of hospitalized patients with COVID-19 analyzed to date, we find that increased immature granulocyte and neutrophil counts early in hospitalization are associated with increased mortality. The presence of conspicuous neutrophil activation in critically ill patients with COVID-19 may not have been detected in prior analyses of circulating biomarkers and flow cytometry because neutrophil granule proteins are not included in standard cytokine panels and because neutrophils tend to be excluded during routine preparations of peripheral blood mononuclear cells. By identifying a consistent signature of neutrophil development and activation that anticipates the onset of critical illness, our findings suggest that neutrophils may play a central role in the pathogenesis of severe COVID-19.

The host response to infection can be understood as a balance between resistance (the ability to eliminate a pathogen) and tolerance (the ability to maintain essential biological functions in the presence of a pathogen).^21^ The immune response is essential for pathogen resistance, but it can also cause costly tissue injury, leading to a failure of tolerance. Neutrophils and macrophages are critical arms of the innate immune system and are often the first responders to infection and injury. However, neutrophils can also cause significant collateral damage to tissues.^22^ While an important role for monocytes and macrophages in severe COVID-19 has been well described,^2^ a potential role for neutrophils has been suggested by the prognostic value of the neutrophil to lymphocyte ratio,^20^ and recent studies have identified a population of immature neutrophils present in the blood of critically ill COVID-19 patients, as well as increased numbers of neutrophils in the bronchoalveolar lavage.^1,3,4,23^ However, it has remained unclear whether there is heightened neutrophil activation in severe COVID-19, and whether the observed innate immune response is a consequence or potential cause of critical illness.

In this study, we first used plasma proteomics and an unbiased machine-learning analysis, which revealed that the factors that best discriminated between critically ill and non-critically ill patients were neutrophil-related proteins. Two members of this neutrophil signature, G-CSF and IL-8, stimulate neutrophil development and chemotaxis and have previously been associated with severe COVID-19.^5^ It was recently reported that IL-8 levels predict survival in a large cohort of hospitalized patients.^5^ The mechanistic significance of these findings, however, was not clear. In this study, together with G-CSF and IL-8, we identify four proteins that are released from neutrophils upon activation, which are among the strongest predictors of critical illness. Elevated HGF levels have previously been reported in severe COVID-19 but were not associated with neutrophil activity, while RETN, LCN2, and MMP8 are all novel markers of critical illness in COVID-19.^24^ Focusing our attention on this neutrophil activation signature, we found that a machine learning model based only on these markers was also able to accurately classify patients according to their disease severity, indicating that neutrophil activation is a consistent feature found in critically ill patients with COVID-19.

We reasoned that if neutrophil activation plays a causal role in the development of critical illness, then evidence of neutrophil activation should be present prior to the onset of critical illness. To address this question, we analyzed plasma collected longitudinally from a separate cohort of hospitalized patients with COVID-19. We found that the neutrophil activation signature was already elevated on day 1 of hospitalization in patients who were not critically ill at that time but would go on to develop critical illness and require transfer to the ICU. Among all patients, day 1 elevations in neutrophil activation markers were also associated with increased mortality during hospitalization. Interestingly, levels of the neutrophil activation markers within each group were relatively stable when sampled over time. These findings indicate that neutrophil activation precedes the onset of critical illness and suggest, 1) that neutrophils may play an essential role in driving critical illness, which could be targeted therapeutically, and 2) that this neutrophil activation signature has the potential to provide a powerful clinical tool for early detection of the need for higher-level care or targeted therapies.

The neutrophil markers we identified suggest a model of neutrophil development and activation in severe COVID-19: We hypothesize that high levels of the growth factor G-CSF stimulate neutrophil production and that the chemokine IL-8 (CXCL8) drives neutrophil migration into the lung and perhaps other tissues.^12,13,25^ Among neutrophils’ key effector mechanisms is the release of granules containing proteins with antimicrobial and other functions in inflammation. RETN, LCN2, HGF, and MMP8 are well-established products of neutrophil secondary granules,^7-10,26,27^ and detection of these proteins at high concentrations in the circulation of critically ill COVID-19 patients strongly suggests neutrophil activation and degranulation. The high degree of correlation between these markers and ANC further supports that model.

Three of these proteins, RETN, LCN2, and MMP8, were also among the top transcriptional markers of a developing neutrophil population that has been identified specifically in the blood of severely ill COVID-19 patients.^3,4^ Although these cells could in principle be the direct source of the proteins we detect in circulation, it is more likely that the genes are transcribed in the developing neutrophils and subsequently packaged into granules and released from mature neutrophils, a sequence of events that is well-described in the neutrophil literature. Constituents of secondary granules, including RETN, LCN2, and MMP8, are known to be transcribed specifically in the myelocyte stage of development by the transcription factor C/EBPε, whose expression is essentially restricted to this phase.^8,16,27-29^ C/EBPε was also highly enriched in the observed developing neutrophil population,^3^ suggesting that these cells are largely myelocytes that have entered the circulation. Typically, neutrophil development takes place in the bone marrow, and immature forms like myelocytes are not detected in the blood. However, during emergency granulopoiesis, neutrophil development accelerates, and immature granulocytes exit the bone marrow and enter the circulation. This process is thought to be driven by high levels of G-CSF, which we detect in the blood of severely ill patients in this study.^12^ Furthermore, in our analysis of a hospital-wide dataset, we find that the earliest counts of immature granulocytes (which include myelocytes) predict mortality. Altogether, a model of emergency granulopoiesis followed by neutrophil activation is beginning to emerge as a key feature of critical illness in COVID-19.

Components of our neutrophil activation signature have effector functions that may be detrimental to patients with COVID-19. RETN regulates production of multiple cytokines, including IL-6, IL-8, and TNF-α,^30^ and is associated with increased endothelial permeability and risk of thrombosis.^31^ LCN2, while initially described to have antimicrobial activities,^32^ is highly upregulated in autoimmune diseases such as systemic lupus erythematosus,^33^ Kawasaki disease,^34^ and inflammatory bowel disease.^35^ Elevated levels of MMP8 were associated with worsening clinical outcomes in pediatric patients with ARDS.^36^ Inappropriate release of these and other components of neutrophil granules may drive key aspects of critical illness in COVID-19, as suggested by previous studies investigating the contribution of neutrophils to acute lung injury.^37^

The signature of neutrophil activation that we identify is accompanied by monocyte and macrophage activation that has previously been described in severe COVID-19. Cytokines produced by both activated neutrophils and macrophages were elevated on day 1 of hospitalization in patients who would ultimately develop critical illness and in patients who did not survive, suggesting that these cell types may function together in the pathogenesis of severe disease. Neutrophil and macrophage responses are highly coordinated with one another in infection and injury. For instance, neutrophil granule proteins stimulate monocyte chemotaxis, macrophages are responsible for phagocytosing apoptotic neutrophils, and macrophage-derived proteins, including IL-8, G-CSF, and TNF, stimulate neutrophil chemotaxis and survival.^38^ Interestingly, whereas macrophage-derived cytokines are also markedly increased in non-ICU patients compared to controls, the release of neutrophil-derived granule proteins appears to be more restricted to patients who are or will become critically ill, suggesting that neutrophil activation and degranulation may be a key event in the progression to severe disease.

Neutrophils classically respond to bacterial infection or tissue injury; whether SARS-CoV-2 induces a specific type of cellular or tissue damage that promotes neutrophil activation will require further investigation. Moreover, the specific cell types that express G-CSF and IL-8 in COVID-19 will need to be elucidated. It is possible that there is a threshold of injury to the lungs, and perhaps the vascular beds, that triggers neutrophil activation,^39,40^ setting in motion the cascade of events that propagate critical illness in COVID-19. This might explain the observation that many patients in the hospital initially appear clinically stable and then rapidly deteriorate, requiring ICU care and mechanical ventilation. Directly testing the causal role of neutrophil activation in COVID-19 will require animal models that recapitulate the activation of neutrophils and allow for manipulation, or targeted therapeutic intervention in patients. However, it is notable that neutropenic patients who have received exogenous G-CSF show increased mortality, providing further support for a potential causal relationship between neutrophil development and activation and adverse clinical outcomes.^41,42^

Our study suggests several potential therapeutic strategies, including inhibition of G-CSF, IL-8, or other drivers of neutrophil activation, but the potential benefit of such strategies will need to be weighed carefully against the risks of modulating a key aspect of the innate immune system. The recent finding that dexamethasone improves mortality in COVID-19 offers strong support for the model that immunopathology plays an important role in COVID-19 pathogenesis,^43^ but it does not identify the key immune components involved in this process. It is notable, however, that one of the oldest recognized effects of corticosteroid treatment is the inhibition of neutrophil binding to endothelial cells, preventing infiltration into tissues (and paradoxically increasing their numbers in circulation).^44^ It will be essential to continue to close the gap in our understanding of how the host immune response determines clinical outcomes in COVID-19 and how this response can be selectively targeted to provide safer and more effective immunomodulatory treatments.

In summary, we demonstrate that increased circulating levels of neutrophil activators (G-CSF, IL-8) and neutrophil effectors (RETN, LCN2, HGF, and MMP8) are hallmarks of critical illness in COVID-19 and that they identify high risk patients upon initial admission to the hospital, prior to the onset of critical illness. We also demonstrate, in one of the largest cohorts of COVID-19 patients to date, that early rises in immature granulocyte and neutrophil counts are associated with increased mortality, suggesting that our mechanistic insights are generalizable to broader COVID-19 patient populations. These findings represent key advances toward understanding the mechanisms of COVID-19 pathogenesis, developing more accurate prognostic indicators, and most importantly, guiding the next generation of therapeutic strategies for COVID-19.

## Materials and Methods

### Study Design and Participants

We first conducted a study of 49 adult patients admitted to Yale-New Haven Hospital between April 13, 2020 and April 24, 2020 with a confirmed diagnosis of COVID-19 via polymerase chain reaction (designated ‘cross-sectional’ cohort). The protocol was approved by the Yale University Institutional Review Board (IRB #2000027792). Plasma samples were analyzed from 40 patients who were treated in a medical ICU and 9 patients in a non-ICU COVID-19 unit in our hospital. Admission to the ICU was based on established ICU admission guidelines (**Supp. Table 1**). For this cohort, blood was collected at a single timepoint during the course of hospitalization (**Supp. Fig. 8**). Blood samples collected from an additional 13 asymptomatic, non-hospitalized, presumed SARS-COV-2 negative controls were also analyzed after signed consents were obtained for a separate approved IRB protocol (IRB #1401013259).

We also analyzed blood samples obtained longitudinally on day 1 (within 24 hours), day 4, and day 7 of hospitalization from a separate cohort of 23 consecutive adult patients who were admitted for treatment of laboratory-confirmed COVID-19 between May 23, 2020 and May 28, 2020 and remained hospitalized until at least day 4 (designated ‘longitudinal’ cohort).

Lastly, we used the Yale Department of Medicine Covid Explorer (DOM-CovX) database to evaluate blood count data from a total of 3,325 deidentified COVID-19-positive patients admitted to the six hospitals within the Yale New Haven Health System (IRB #2000028509) (designated ‘CovX’ cohort). Patients with a confirmed positive COVID-19 test within 14 days preceding their hospitalization were included in the cohort. This dataset combines all clinical variables extracted from the electronic medical record (Epic, Verona WI) including demographics, comorbidities, procedures, and all laboratory recorded during the hospitalization.

#### Procedures

Blood was collected in 3.2% sodium citrate tubes and centrifuged at 2000 g for 20 minutes at room temperature, and the resulting plasma supernatant was frozen at −80°C and used for further testing. The biomarker profiling analyses were conducted at Eve Technologies (Calgary, Alberta, Canada). For the cross-sectional cohort, the following assays were performed: Human Cytokine 71-Plex, Human Complement Panels 1 and 2, Human SAA & ADAMTS13, and Human Adipokine 5-Plex. For the longitudinal cohort, the following assays were performed: Human Cytokine 48-Plex, Human Complement Panel 1, Human Adipokine 5-Plex, and Human MMP 9-Plex and TIMP 4-Plex. Five of the control samples were evaluated concurrently with the longitudinal cohort samples (**Table 1b**). Heatmaps were generated using the concentrations of circulating biomarkers obtained from biomarker profiling analyses using Heatmapper as described.^45^

#### Principal component analysis and random forest classifier

Values for biomarker profiles of patients in the cross-sectional cohort were log transformed. A pseudocount of half the minimum observed non-zero value per biomarker was added to each observed zero value before log 10 transformation. IL-6 was excluded from modeling, as most subjects (38 out of 40) in the ICU subset of the cross-sectional cohort had received IL-6 receptor blockade prior to blood collection. The remaining biomarker values were used in a principal component analysis and a random forest classifier using the scikit-learn python package.^46^ Data were partitioned into 66-33 train-validation split (training cohort: 26 ICU patients, 7 non-ICU patients, 3 controls; validation cohort: 14 ICU patients, 2 non-ICU patients, 5 controls), and all biomarkers were then used to predict the ICU status. A maximal tree depth of 10 was used and the minimal cost-complexity was set to 0.02, otherwise all other parameters were set to their defaults. Feature importance was assessed using mean decrease in impurity. Data restricted to five biomarkers of interest with high feature importance were then used in a separate random forest classifier, using the same methods.

#### Single cell analyses

Processed count matrices with de-identified metadata and embeddings from a single-cell RNAseq dataset published by Wilk et al were downloaded from the COVID-19 Cell Atlas (https://www.covid19cellatlas.org/#2iki20) hosted by the Wellcome Sanger Institute.^3^ This dataset was further analyzed using Seurat (version 3.0, https://satijalab.org/seurat). The clusters in this object were renamed based on cell type, using the UMAPs and heat maps provided. A new metadata column, “ARDSstatus” was added to the object, which grouped the samples into the categories “Control”, “COVID19-non-ARDS”, and “COVID19-ARDS”. Violin and Feature Plots were generated to examine specific genes of interest. The cells identified as “Developing Neutrophil’’ were subsetted from the overall object to examine this population more closely.

#### Statistical Analyses

To evaluate differences in mean age across the groups we used one-way ANOVA with post-hoc Tukey’s multiple comparisons tests in the cross-sectional cohort and Kruskal-Wallis test in the longitudinal cohort, as the samples in the former cohort but not the latter conformed to the normal distribution. For the comparisons of proportions of men and women in the cohorts we used Chi-square tests in the cross-sectional cohort and Fisher’s exact test in the longitudinal cohort. In both cross-sectional and longitudinal cohorts, we examined differences in proportions of patients with comorbidities using Fisher’s exact test.

To compare biomarker values, we used two-sample tests. As most sample distributions did not satisfy Anderson-Darling and D’Agostino-Pearson tests of normality, we used the unpaired two-tailed Mann-Whitney u-test. For samples that conformed to the normal distribution we used unpaired two-sided t-tests with Welch correction for unequal variances, where applicable; where appropriate, *p* values were corrected for multiple comparisons using the false discovery rate procedure with the discovery rate Q of 5%. *P* values of less than 0.05 were considered significant.

Correlation coefficients between absolute neutrophil count and each biomarker was computed using Spearman’s rank method. For Kaplan-Meier survival analyses we used median values of evaluated variables as cut points to classify patients into two groups, high and low, based on whether patients’ values for a particular biomarker or cell count were above or below the cut point. We then used logrank test to compare the two groups of patients for each evaluated variable. To determine p-values for cell-type-specific enrichment of genes in single-cell RNAseq data, we used Wilcoxon Rank Sum test. All statistical analyses were carried out using GraphPad Prism (v8.4.3, GraphPad Software, San Diego, CA), Stata (v16, StataCorp. College Station, TX), and R (v4, R Core Team, 2020).

## Data Availability

N/A

## Acknowledgments

This work was supported by a gift donation from Jack Levin to the Benign Hematology program at Yale, the National Institutes of Health (HL142818 to HJC, DK079310 and DK113191 to FPW, GM136651 and HL139116 to MLM), the American Heart Association (Transformational Project Award to HJC), and the DeLuca Center for Innovation in Hematology Research at Yale Cancer Center. We would like to thank all front-line staff taking care of our community of patients affected by SARS-COV-2 and are grateful for the multidisciplinary collaboration at the Yale School of Medicine and Yale-New Haven Health System in the care of our community.

## Notes

### Competing Interest Statement

The authors have declared no competing interest.

### Author Declarations

Yale University approved IRB #s:2000027792, 1401013259 and 2000028509

## References

1. Liao, M., et al. Single-cell landscape of bronchoalveolar immune cells in patients with COVID-19. Nat Med 26, 842–844 (2020).

2. Merad, M. & Martin, J.C. Pathological inflammation in patients with COVID-19: a key role for monocytes and macrophages. Nat Rev Immunol 20, 355–362 (2020).

3. Wilk, A.J., et al. A single-cell atlas of the peripheral immune response in patients with severe COVID-19. Nat Med (2020).

4. Schulte-Schrepping, J., et al. Suppressive myeloid cells are a hallmark of severe COVID-19. *medRxiv*, 2020.2006.2003.20119818 (2020).

5. Del Valle, D.M., et al. An inflammatory cytokine signature predicts COVID-19 severity and survival. Nat Med (2020).

6. Jiang, S., et al. Human resistin promotes neutrophil proinflammatory activation and neutrophil extracellular trap formation and increases severity of acute lung injury. J Immunol 192, 4795–4803 (2014).

7. Bostrom, E.A., Tarkowski, A. & Bokarewa, M. Resistin is stored in neutrophil granules being released upon challenge with inflammatory stimuli. Biochim Biophys Acta 1793, 1894–1900 (2009).

8. Chumakov, A.M., Kubota, T., Walter, S. & Koeffler, H.P. Identification of murine and human XCP1 genes as C/EBP-epsilon-dependent members of FIZZ/Resistin gene family. Oncogene 23, 3414–3425 (2004).

9. Crestani, B., et al. Differential role of neutrophils and alveolar macrophages in hepatocyte growth factor production in pulmonary fibrosis. Lab Invest 82, 1015–1022 (2002).

10. Grenier, A., et al. Presence of a mobilizable intracellular pool of hepatocyte growth factor in human polymorphonuclear neutrophils. Blood 99, 2997–3004 (2002).

11. Bundgaard, J.R., Sengelov, H., Borregaard, N. & Kjeldsen, L. Molecular cloning and expression of a cDNA encoding NGAL: a lipocalin expressed in human neutrophils. Biochem Biophys Res Commun 202, 1468–1475 (1994).

12. Manz, M.G. & Boettcher, S. Emergency granulopoiesis. Nat Rev Immunol 14, 302–314 (2014).

13. Kolaczkowska, E. & Kubes, P. Neutrophil recruitment and function in health and inflammation. Nat Rev Immunol 13, 159–175 (2013).

14. Vabret, N., et al. Immunology of COVID-19: Current State of the Science. Immunity 52, 910–941 (2020).

15. Huang, C., et al. Clinical features of patients infected with 2019 novel coronavirus in Wuhan, China. Lancet 395, 497–506 (2020).

16. Lawrence, S.M., Corriden, R. & Nizet, V. The Ontogeny of a Neutrophil: Mechanisms of Granulopoiesis and Homeostasis. Microbiol Mol Biol Rev 82(2018).

17. Hasty, K.A., et al. Human neutrophil collagenase. A distinct gene product with homology to other matrix metalloproteinases. J Biol Chem 265, 11421–11424 (1990).

18. Dorweiler, B., et al. Subendothelial infiltration of neutrophil granulocytes and liberation of matrix-destabilizing enzymes in an experimental model of human neo-intima. Thromb Haemost 99, 373–381 (2008).

19. Khanna-Gupta, A., et al. Human neutrophil collagenase expression is C/EBP-dependent during myeloid development. Exp Hematol 33, 42–52 (2005).

20. Liu, Y., et al. Neutrophil-to-lymphocyte ratio as an independent risk factor for mortality in hospitalized patients with COVID-19. Journal of Infection (2020).

21. Medzhitov, R., Schneider, D.S. & Soares, M.P. Disease Tolerance as a Defense Strategy. Science 335, 936–941 (2012).

22. Bardoel, B.W., Kenny, E.F., Sollberger, G. & Zychlinsky, A. The balancing act of neutrophils. Cell Host Microbe 15, 526–536 (2014).

23. Zhou, Z., et al. Heightened Innate Immune Responses in the Respiratory Tract of COVID-19 Patients. Cell Host Microbe 27, 883-890.e882 (2020).

24. Liu, Y., et al. Elevated plasma levels of selective cytokines in COVID-19 patients reflect viral load and lung injury. National Science Review 7, 1003–1011 (2020).

25. Lacy, P. Mechanisms of degranulation in neutrophils. Allergy Asthma Clin Immunol 2, 98–108 (2006).

26. Cramer, E.P., et al. Lipocalin-2 from both myeloid cells and the epithelium combats Klebsiella pneumoniae lung infection in mice. Blood 129, 2813–2817 (2017).

27. Serwas, N.K., et al. CEBPE-Mutant Specific Granule Deficiency Correlates With Aberrant Granule Organization and Substantial Proteome Alterations in Neutrophils. Frontiers in Immunology 9(2018).

28. Cowland, J.B. & Borregaard, N. Granulopoiesis and granules of human neutrophils. Immunological Reviews 273, 11–28 (2016).

29. Gombart, A.F., et al. Regulation of neutrophil and eosinophil secondary granule gene expression by transcription factors C/EBPε and PU.1. Blood 101, 3265–3273 (2003).

30. Zhang, Z., et al. Resistin induces expression of proinflammatory cytokines and chemokines in human articular chondrocytes via transcription and messenger RNA stabilization. Arthritis Rheum 62, 1993–2003 (2010).

31. Jamaluddin, M.S., Weakley, S.M., Yao, Q. & Chen, C. Resistin: functional roles and therapeutic considerations for cardiovascular disease. Br J Pharmacol 165, 622–632 (2012).

32. Devireddy, L.R., Hart, D.O., Goetz, D.H. & Green, M.R. A mammalian siderophore synthesized by an enzyme with a bacterial homolog involved in enterobactin production. Cell 141, 1006–1017 (2010).

33. Rubinstein, T., Pitashny, M. & Putterman, C. The novel role of neutrophil gelatinase-B associated lipocalin (NGAL)/Lipocalin-2 as a biomarker for lupus nephritis. Autoimmun Rev 7, 229–234 (2008).

34. Biezeveld, M.H., et al. Sustained activation of neutrophils in the course of Kawasaki disease: an association with matrix metalloproteinases. Clin Exp Immunol 141, 183–188 (2005).

35. Nielsen, B.S., et al. Induction of NGAL synthesis in epithelial cells of human colorectal neoplasia and inflammatory bowel diseases. Gut 38, 414–420 (1996).

36. Kong, M.Y., Li, Y., Oster, R., Gaggar, A. & Clancy, J.P. Early elevation of matrix metalloproteinase-8 and -9 in pediatric ARDS is associated with an increased risk of prolonged mechanical ventilation. PLoS One 6, e22596 (2011).

37. Grommes, J. & Soehnlein, O. Contribution of neutrophils to acute lung injury. Mol Med 17, 293-307 (2011).

38. Prame Kumar, K., Nicholls, A.J. & Wong, C.H.Y. Partners in crime: neutrophils and monocytes/macrophages in inflammation and disease. Cell and Tissue Research 371, 551–565 (2018).

39. Goshua, G., et al. Endotheliopathy in COVID-19-associated coagulopathy: evidence from a single-centre, cross-sectional study. Lancet Haematol (2020).

40. Varga, Z., et al. Endothelial cell infection and endotheliitis in COVID-19. Lancet 395, 1417–1418 (2020).

41. Morjaria, S., et al. The Effect of Neutropenia and Filgrastim (G-CSF) in Cancer Patients With COVID-19 Infection. *medRxiv*, 2020.2008.2013.20174565 (2020).

42. Nawar, T., et al. Granulocyte-colony stimulating factor in COVID-19: Is it stimulating more than just the bone marrow? American Journal of Hematology 95, E210-E213 (2020).

43. Group, R.C., et al. Dexamethasone in Hospitalized Patients with Covid-19 - Preliminary Report. N Engl J Med (2020).

44. Nakagawa, M., et al. Glucocorticoid-induced granulocytosis: contribution of marrow release and demargination of intravascular granulocytes. Circulation 98, 2307–2313 (1998).

45. Babicki, S., et al. Heatmapper: web-enabled heat mapping for all. Nucleic Acids Res 44, W147-153 (2016).

46. Hazan, H., et al. BindsNET: A Machine Learning-Oriented Spiking Neural Networks Library in Python. Front Neuroinform 12, 89 (2018).

